# A protocol for the comparison of telephone and in-person interview modalities: duration, richness, and costs in the context of exploring determinants of equitable access to community health services in Meru, Kenya

**DOI:** 10.1101/2024.03.04.24303701

**Authors:** Luke N Allen, Sarah Karanja, Malebogo Tlhajoane, John Tlhakanelo, David Macleod, Andrew Bastawrous

**Author notes:** **Corresponding author:**, Department of Clinical Research, Faculty of Infectious and Tropical Diseases, London School of Hygiene and Tropical Medicine, Keppel Street, London, WC1E 7HT. **Author contributions:** LA conceived the study and designed the methods with SK. LA wrote the initial draft. SK and MT provided additional written content. JT provided health economics input and reviewed the protocol. DM provided statistical input and reviewed the protocol. AB reviewed the protocol and provided additional methodological comments. All authors reviewed and approved the final version of the protocol.

## Abstract

**Background:** Our research team is conducting phenomenological interviews with people who have not been able to access health services in Meru County, Kenya, aiming to explore the barriers they face and their perceptions of how we could modify our community outreach services to improve accessibility. We plan to conduct an embedded study that compares in-person and telephone interview modalities in terms of the richness of the data and the resources required for each modality.

**Methods/Design:** This is a qualitative mode comparison study, embedded within a broader project to understand and address the issues that lead to inequitable access to local outreach clinics in Kenya. We will recruit at least 40 people who have been referred to local services but who have not been able to attend. We will conduct in-person interviews with half of these people, and telephone interviews with the other half. We will use random numbers to determine the modality that is used for each participant. All interviews will be conducted in the same month by a team of six research assistants who will use the same topic guide and analytic matrix for each interview. For all interviews conducted in each mode we will record and compare the mean duration; mean number of themes reported by each participant; total number of themes reported; interviewer rating of perceived richness; interviewer rating of perceived ease of building rapport; number of days taken by the team to complete all interviews; and all costs associated with conducting the interviews.

**Discussion:** The findings will help us to weigh up the relative strengths and weaknesses of each modality for our research context. Given that we are exploring a focused research question in a fairly homogenous population, we anticipate that there may not be a meaningful difference in the number of themes reported.

## 1. Background

### 1.1 Telephone and in-person interviews

Our research team is using interviews to explore barriers to health service access and potential solutions by engaging with people who were referred to local eye clinics but were not able to access care.(Allen & et al, 2023) Our project is embedded within Kenya’s ‘Vision Impact Project’ (VIP) eye screening programme that operates in ten counties. Over a million people have been screened in the past year, and over 150,000 of these people have received care in free local outreach clinics (Peek Vision, 2018). However, internal data from the screening programme suggest that up to half of all those referred to local treatment clinics are not able to access care. Furthermore, early data from a related study suggests that certain sociodemographic groups have much lower odds of accessing care than others. In Meru County we have found that younger adults (aged 18-44 years) are the least likely to receive the care that they need. We want to explore these peoples’ experiences and perceptions of specific barriers to accessing care, as well as their ideas around any changes we could make to the eye care services to make it easier to access care. Qualitative interviews – especially those grounded in the phenomenological approach - are designed to elicit rich data about participants’ lived experiences and perceptions of a given phenomenon (Pope & Mays, 2020).

The VIP screening budget is limited, and programme implementers are keen that our interviews can be conducted quickly and as inexpensively as possible whilst still delivering robust findings. The incentive to keep time and costs low is further underlined by our desire to see embedded qualitative research adopted more widely across routine programmatic quality improvement initiatives, so that the voices of intended service beneficiaries can be included in decision making. Based on the findings of a recent scoping review on rapid qualitative research methods (Allen et al., 2023) we have developed a rapid and ‘abductive’ interview approach that uses a deductive analytic matrix to facilitate rapid iterative analysis of data whilst “making space for inductive identification of themes and issues not predicted at the outset”(Pope & Mays, 2020). Our work employs a phenomenological approach, grounded in a pragmatist philosophical paradigm.

The work in Kenya is part of a broader overall project to develop equity-driven and evidence-based approaches to improve access to community-based services across Kenya, Botswana, India and Nepal (Allen & et al, 2023). Hundreds of thousands of people are being screened and referred to local services each year, however only around half are able to access care. We want to develop an interview approach that can be taken to scale across these four countries – and potentially beyond – to deliver timely insights into how these programmes can be made more accessible, especially for ‘left behind’ groups. Given the scale of the project, telephone interviews are likely to offer the most pragmatic means of obtaining timely insights on how to improve services, however it is not clear what – if anything - would be lost.

### 1.2 Comparing mode effects

In-person interviews are commonly perceived the ‘best’ way of obtaining rich phenomenological data due to the fact that the interviewer can observe visual cues and potentially build rapport more easily (Novick, 2008; Rahman, 2023; Rubin & Rubin, 2011). However, telephone interviews offer unique advantages: the increased social distance can make it easier for participants to discuss sensitive topics, travel time and interviewer safety concerns are eliminated, power imbalances are partially concealed, and overall costs can be greatly reduced – depending on the specific study design and population (Novick, 2008; Sturges & Hanrahan, 2004; Vogl, 2013). Given that our participants are spread across vast distances, the risks, costs, and time-requirements for in-person interviews are likely to compare poorly with telephone interviewing.

## 2. Methods

### 2.1 Aims and hypothesis

In this study, we aim to compare in-person and telephone interviews in terms of data richness, time taken to complete a comparable number of interviews, and associated costs. We hypothesise that telephone interviews will be shorter and cheaper than in-person interviews, but offer less rich data.

### 2.2 Research question

Which modality offers the best balance of richness, duration, and costs in the context of an embedded study to explore barriers and solutions to low access to care in Meru County, Kenya? Note that we aim to deliver timely and robust findings at the lowest possible cost.

#### 2.2.1 Setting and participants

Interviews will be conducted with people who were referred to local treatment outreach clinics in Meru County as part of the VIP screening programme, but who were not able to access care.

### 2.3 Sampling and recruitment

We will obtain a list of all those who did not attend clinic within two weeks of their appointed data from Peek Vision, a partner organisation that provides the screening and patient flow management software for the programme. Peek also record contact numbers for all participants. We will generate a list of people belonging to the sociodemographic groups with the lowest overall odds of accessing care; people aged 18-44 years old. We will sample from this list, using computer-generated random numbers to identify interviewees and determine interview modality. Whilst a standard qualitative approach would use saturation to determine sample size, we want to ensure that we have a sufficient number of interviews to enable comparison between both modalities. Empirical research has found that thematic saturation is often reached within 9-17 interviews, given a relatively homogenous population.(Francis et al., 2010; Guest et al., 2006) We will budget for at least 40 interviews; 20 in-person and 20 via telephone.

We will call potential participants to explain the study and invite them to participate. The interview modality for each potential participant will be decided before the recruitment call is placed, based on random numbers i.e. participants will not be able to choose their modality. For those who agree to an in-person interview, the researchers will then arrange a time to visit in-person. For those who agree to a telephone interview, the researchers will either proceed with the interview or agree on a more convenient time to call back. Recorded verbal informed consent will be sought for telephone interviews and written informed consent will be sought for in-person interviews.

### 2.4 Study design

We will compare an equal number of interviews conducted using each modality, with a minimum of 20 v 20. The same team of data collectors will conduct all interviews using the same semi structured interview guide for both modalities. The same process for audio recording data and directly transcribing quotes into the deductive analytic matrix will be used for both modalities, and the same process of iterative review and analysis across all cases within each modality will be used to generate the final themes.

### 2.5 Domains

#### 2.5.1 Interview duration

We will measure the duration of each interview from the start of the consenting process until the researcher concludes the interview e.g. by thanking the participant for answering all of their questions. This will be used as a proxy for richness, based on the assumption that longer interviews capture richer data than shorter interviews, as reported in the methods literature (Irvine et al., 2013; Johnson et al., 2021; Sturges & Hanrahan, 2004; Vogl, 2013). We will report the mean interview duration and the range for each modality.

#### 2.5.2 Matrix wordcount

We will count the total number of words entered into the analytic matrix. These are all verbatim quotes directly transcribed from the audio by the researchers. Following previous studies, we will assume that a higher wordcount is associated with richer data (Johnson et al., 2021; Sturges & Hanrahan, 2004).

#### 2.5.2 Total number of themes

Following the approach used by Abrams et al. and Johnson et al, we will report the aggregated total number of unique themes, reported separately for barriers and solutions, that are reported across all interviews conducted using each modality, assuming that the modality that captures the largest number of unique themes is capturing richer data (Abrams et al., 2015; Johnson et al., 2021). From an operational standpoint, our underlying study is primarily concerned with generating potential solutions that will improve equitable access, so the number of unique solutions that emerge from each set of interviews is a particularly important metric.

#### 2.5.3 Number of themes reported by each participant

We will also report the range and mean number of unique themes (barriers and solutions) identified by each participant for each modality. This is to hedge against a situation where one modality generates a greater number of themes than the other but this is driven by one individual.

#### 2.5.4 Interviewer subjective rating of richness

After all interviews are complete, each of the six data collectors will provide a single global summary rating of the perceived richness obtained from all in-person and all telephone interviews, using a simple Likert scale: low = 1, moderate = 2, high = 3, after the approach used in two previous studies (Abrams et al., 2015; Johnson et al., 2021).

#### 2.5.5 Interviewer subjective rating of rapport

Again, after all interviews are complete, each researcher will provide a global summary rating of the perceived ease of building rapport across all in-person and all telephone interviews using a simple Likert scale: low = 1, moderate = 2, high = 3.

#### 2.5.6 Time taken to plan and complete all interviews

We will report the total amount of time taken to plan and complete all interviews in each modality to the nearest half-day. We anticipate that in-person interviews will take longer due to the planning and logistics requirements.

#### 2.5.7 Costs

We will record costs from the payer’s perspective. Both modalities use the same sampling and analytic approach, so we will only compare costs associated with data collection. For telephone interviews these costs include airtime and staff daily salaries multiplied by the number of days taken for data collection, starting with the first phone call to recruit the first participant and ending with the conclusion of the final interview. Voice recorders purchased for telephone interviews will also be used to record in-person interviews, so these will not be included. Similarly, the two-day training of data collectors that we delivered included training for both modalities, so this cost will not be included.

For in-person interviews we will include the costs of printing consent forms, transport for researchers, transport reimbursement offered to participants; incentives to local Community Health Promoters and sub-county health officials to assist with setting up the interviews (mobilisation/sensitisation), and staff daily salaries multiplied by the number of days taken for data collection.

We will not compare overhead costs unless they differ for the modalities. The local research manager will record any unforeseen additional costs associated with each modality.

#### 2.5.8 Triangulation of themes

We will compare the themes that emerge from both modalities; identifying those that are identified by both modalities (agreement); one but not the other (silence); and any areas of dissonance. An example of dissonance would be one set of interviews identifying a barrier around spectacles being perceived to be too expensive, whilst the other identifies a barrier around the perceived cheapness of spectacles inferring poor quality.

### 2.7 Statistical approach

For quantitative comparisons we will use histograms to check the data for normality and then use either unpaired T-tests or Mann-Whitney tests, as appropriate, to provide evidence as to whether the two modalities differ in any of the domains

## 3. Ethics

Ethical approval has already been granted by the Kenya Medical Research Institute (KEMRI), the Kenyan National Commission For Science, Technology & Innovation (NACOSTI), and the London School of Hygiene & Tropical Medicine research ethics committee. Each participant will be provided with information that described the aims and purposes of the study, and asked to provide written or verbal consent, depending on the modality. All consenting and interviews will be conducted in the participant’s preferred language. All interviews will be audio recorded and we anticipate that they will last approximately 20 minutes. There are negligible risks involved, however, should a participant become distressed during the interview, the researcher will provide appropriate support. The nature of the topics will be discussed before the interview commences and the participants will be free to terminate the interview at any time without providing a reason. Financial reimbursement will be provided to those who incur travel costs to participate in in-person interviews e.g. if they are not conducted in their homes.

## 4. Discussion

Many different proxies and domains have been used in previous mode-comparison research. Our study uses eight different domains, covering costs, time, and six proxies for richness. We will not use word count or interviewer dominance measures as these both require typed transcripts, and our approach is based around direct data entry from recorded audio.

The findings of this study will help us to weigh up the relative strengths and weaknesses of each mode for our purpose and context. Given that we are exploring a relatively focused research question in a fairly homogenous population, we anticipate that we will find that the richness of the data obtained will be roughly equivalent. This raises an important question; how should we balance differences in richness, costs, and time requirements? At what threshold would pragmatic advantages outweigh putative methodological benefits? There is not a simple answer to this question. We plan to report the performance of each modality across each of our domains and come to a holistic judgement.

By presenting data on the performance of each mode across a range of domains we hope that our findings will be of use to a wider body of researchers who will be able to apply their own weightings and come to their own conclusions about which mode would be preferable for their given context. For our own broader research team, we will use these findings to decide whether to pursue in-person interviews in Botswana, India, and Nepal.

A major strength of this study will be that it uses multiple measures of data richness. A key limitation is that our study is designed to generate evidence of superiority rather than equivalence. Generating evidence that the two approaches are ‘significantly’ different from one another would likely require a larger sample size than our underlying study will generate.

## Ethical approval

The study was granted ethical approval by KEMRI scientific and ethics review unit on November 30^th^ 2022, NACOSTI on 12^th^ January 2023, and the LSHTM ethics committee on 5^th^ May 2023.

## Data Availability

Data produced in the present study are available upon reasonable request to the authors

